# Short-Term Effects of Strengthening Exercises of Lower Limb Rehabilitation Protocol on Pain, Stiffness, Physical Function and Body Mass Index among Knee Osteoarthritis Participants Who Were Overweight or Obese: A Clinical Trial

**DOI:** 10.1101/2021.11.20.21266300

**Authors:** Muhammad Tariq Rafiq, Mohamad Shariff Abdul Hamid, Eliza Hafiz

**Affiliations:** Centre for Sport and Exercise Sciences, University of Malaya, 50603 Kuala Lumpur, Malaysia; Rehmatul-Lil-Alameen Postgraduate Institute of Cardiology, Punjab Employees Social Security Institution, Lahore, Pakistan; Faculty of Medicine, Dean’s Office, University of Malaya, 50603 Kuala Lumpur, Malaysia

## Abstract

**Background:** Osteoarthritis (OA) of the knee is defined as a progressive disease of the synovial joints and is characterized by wear and tear of cartilage and underlying bone. This study aimed to determine the short-term effects of the lower limb rehabilitation protocol (LLRP) on pain, stiffness, physical function, and body mass index (BMI) among knee OA participants who were overweight or obese.

**Methodology:** Single blinded randomized controlled trial of one-month duration was conducted at Rehmatul-Lil-Alameen Postgraduate Institute, Lahore, Pakistan. Fifty overweight or obese participants with knee OA were randomly divided into two groups by a computer-generated number. Participants in the Rehabilitation Protocol Group (RPG) were provided with leaflets explaining the strengthening exercises of the LLRP and instruction of daily care (IDC), while the participants in the Control Group (CG) were provided with leaflets explaining IDC only for a duration of four weeks. The primary outcome measures were the Western Ontario and McMaster Universities Osteoarthritis Index (WOMAC) scores for pain, stiffness, and physical function. The secondary outcome measures were BMI, exercise adherence, and patients’ satisfaction by the numeric rating scale ranging from 0 to 10. Paired Samples t-test was used to analyze the differences within groups from baseline to post-test evaluations. The analysis of variance 2 × 2 factor was used to analyze the difference of BMI, knee pain, stiffness, and physical function between the groups.

**Results:** Participants in the RPG and CG reported a statistically significant reduction in knee pain, and stiffness (p ≤ 0.05) within group. The reduction in the scores of knee pain was higher in participants of the RPG than the CG (p = 0.001). Additionally, participants in the RPG reported greater satisfaction (p = 0.001), higher self-reported exercise adherence (p = 0.010) and coordinator-reported exercise adherence (p = 0.046) compared to the participants in the CG.

**Conclusion:** Short-term effects of the LLRP appear to reduce knee pain and stiffness only, but not physical function and BMI.

**TRIAL REGISTRATION:** **Name:** Iranian Registry of Clinical Trials

**Number:** IRCT20191221045846N2

**Date of registration:** June 28, 2020

**Registration timing**: prospective

## 1. Introduction

The Knee joint is a complex synovial joint in the human body where the femur, tibia, fibula, and patella articulate [1]. Articulation is supported by structures that include muscles, ligaments, tendons, articular cartilage, synovial membrane, synovial capsule, meniscus, and fat pad [2]. The synovial fluid and articular cartilage lubricates the knee allowing low-friction joint movement [3]. The articular cartilage protects the subchondral bone from local stresses because of its strong reported excessive joint load in the knee osteoarthritic (OA) [4]. A study provides evidence on the fact that excessive joint load in knee OA patients can lead to an increased inflammatory response, joint pain, and swelling [5]. A recent study compared the body mass index (BMI) of obese and non-obese Knee OA elderly individuals and reported that obesity resulted in less functional mobility, slower gait speed, higher pain intensity and difficulty in performing daily living activities than non-obese [6]. It was excavated that obesity increases the joint load in knee OA patients, resulting in lowering functional mobility and increasing pain intensity. Over the past decade, the prevalence rate of overweight and obesity has increased in the United States of America (USA), Canada, Mexico, France, and Switzerland [7]. However, the prevalence of obesity solely among men and women of the USA population was 37.9 and 41.1%, respectively [8]. A recent retrospective study investigated the physical and functional characteristics of 320 patients with knee OA, reporting that obesity and advanced age were associated with an increased risk of knee OA [9]. Yet another study in the perspective under discussion reached the conclusion that overweight and obesity had a negative impact in increasing pain perception among patients with OA [10].

A study suggested that reduced weight may be related to less demand on the proximal muscles, the internal medial rotator muscle of the knee, to provide stability at the toeLoff during excessive rearfoot motion [11]. A recent exposition led to the fact that by acquiring the habit of regular exercise, knee OA patients can reduce pain, improve quality of life and physical activity. One of the defense mechanisms of the knee, such as body weight control, is useful for healthy aging [9]. All international clinical practice guidelines recommend patients with knee OA, who are overweight or obese, to lose their weight. Adherence may refer to different things and can be used to evaluate the attendance, technique or accuracy of exercise protocols in supervised appointments [12]. Research, in the context of adherence refers to the accomplishment of self-reported and coordinator-reported adherence by the intervention groups.

Clinical guidelines recommend exercise therapy as the primary non-pharmacologic treatment for Knee OA [13]. Because of remarkable evidence demonstrating the beneficial effects of physical exercise among patients with OA, exercise is often indicated as one of the main components in the rehabilitation process [14, 15]. Among the several types of physical exercise programs, muscle strengthening is important because of the relationship between muscle weakness, pain, and mal-function [16, 17]. A current systematic review on non-pharmacological interventions for treating symptoms of knee OA in overweight or obese patients resulted that the most effective intervention that showed improvement of knee pain and function was strengthening exercise. Similarly, it also reported that the combination of diet and exercise was found effective in reducing weight and improving knee pain [18]. A study explained that progressive resistance strength training increases load gradually over the training course to strengthen the major muscle groups and has been recommended to prevent or reduce late-life disability for older adults [19]. Effectiveness of rehabilitation in non-weight-bearing positions as well as strengthening exercises of major muscle groups of the lower limbs may provide more objective data than the standard rehabilitation approaches we are using today to treat overweight and obese knee OA patients. However, there is a gap in knowledge regarding whether strengthening exercises of major muscle groups of lower limbs in non-weight-bearing positions can improve the effects of rehabilitation among overweight or obese knee OA participants. Hence, the current randomized controlled trial aimed to determine the short-term effects of strengthening exercise of the lower limb rehabilitation protocol (LLRP) in non-weight-bearing positions on knee pain, stiffness, physical function, BMI, patient, s satisfaction and exercise adherence in overweight or obese knee OA participants.

The current study aimed to determine the short-term effects of strengthening exercise of the lower limb rehabilitation protocol (LLRP) in nonweight bearing positions on pain, stiffness, physical function, BMI, patient, s satisfaction and exercise adherence in overweight or obese knee OA participants.

## 2. Methodology

### 2.1. Design and Setting

This is a single blinded randomized controlled trial that involved participants diagnosed with knee OA who are overweight or obese. Participants were randomized into the Rehabilitation Protocol Group (RPG), and the Control Group (CG) using a computer-generated simple randomization technique. The study was conducted at the Teaching Bay of Rehmatul-Lil-Alameen Postgraduate Institute of Cardiology (RAIC), Punjab Employees Social Security Institution (PESSI), Lahore, Pakistan. All participants were asked to complete the clinical research form (CRF) following randomization. The CRF gathered sociodemographic information, symptoms of knee pain, stiffness, physical function scores, and BMI. The participant’s satisfaction and exercise adherence were collected after 4-weeks of intervention.

Participants in each treatment group were provided with necessary details about their intervention protocol after randomization. Making explanation of the purpose and constraints of the study, the participants were asked to surrender written informed consent for their participation in the study. All participants were also given a diary and asked to record the attendance of completion their interventions based on leaflets. The current study was approved by the ethical committee of Rehmatul-Lil-Alameen Postgraduate Institute of Cardiology, Punjab Employees Social Security Institution with reference number RAIC PESSI/Estt/2020/36 on date 20-05-2020 and the trial was registered in the Iranian Registry of Clinical Trials with reference number IRCT20191221045846N2 on date 28-06-2020.

### 2.2 Study Participants, Recruitment and Selection

Participants with knee OA who were overweight or obese from the urban community of Punjab, Lahore, Pakistan, were screened. The sample included males and females having OA on one or both knees confirmed by a medical specialist according to the Kellgren and Lawrence radiographic scale for the assessment of OA [20]. The anteroposterior and lateral view of plain radiography of the affected knee/knees were performed in the standing position at the Al - Rehmat Trust Hospital, Lahore.

Participant inclusion criteria were:

i. Aged between 45 – 60 years
ii. Having minimum qualification of matriculation
iii. History of knee pain for more than three months
iv. Overweight (BMI ≥ 25kg/m^2^) or obese (BMI > 30kg/m^2^) [21]
v. Diagnosed with mild or moderate knee OA according to Kellgren and Lawrence radiographic scale [20]

Participant exclusion criteria were:

i. Diagnosed with rheumatoid arthritis, systemic lupus erythematosus, flat foot/feet, or spinal deformities
ii. History of metabolic, hormonal, orthopaedic, or cardiovascular disease
iii. Previous surgery of the knee/s [22]
iv. Inability to be unable to walk independently
v. Injection of knee/s for the last six months [22]

All information related to inclusion and exclusion criteria was gathered from the predefined questionnaire. The researcher recruited the participants by active recruitment strategies such as urban political and welfare organizations via word of mouth by the convenience sampling technique. The list of participants with knee OA in the studied area was obtained from the Welfare Organization by explaining the benefits of study participation. Two study coordinators prepared the list of potential participants of knee OA in the recruitment area. After obtaining the list of potential participants of knee OA, the researcher arranged a meeting with the knee OA participants by calling them on the phone. The meeting was held at the teaching bay of RAIC, PESSI, Lahore, Pakistan, in the presence of a medical specialist. Participants were screened for eligibility to participate in the study. Only participants fulfilling the inclusion and exclusion criteria of the study were invited to participate in this study.

### 2.3. Sample Size

Sample size estimation was performed using the G* Power 3.1.3 software. By assuming the medium effect size f = 0.70, setting α = 0.05, power (1-B) = 0.80, number of groups = 2, number of measurements = 2, the total sample size estimated was 33 participants. After considering the apprehension of drop-out or research mortality, the sample size of 50 participants for the two groups was decided.

### 2.4. Blinding and Allocation

The principle investigator was not blinded in the study. The participants receiving the intervention were kept blinded by simply not informing them of their treatment allocation. The coordinators collecting data were independent individuals from the trials and were unaware of the group allocation. There were different coordinators at the baseline and post-test evaluation. Individuals performing the statistical analysis were kept blinded by labelling the groups with nonidentifying terms (such as X and Y).

### 2.5. Study Randomization

After completing the screening of Knee OA participants, the researcher allocated the 50 selected participants into two groups, namely, RPG and CG, by a computer-generated number. Each group consisted of 25 participants. The participants receiving the intervention were blinded by their treatment allocation. The participants in the RPG followed the strengthening exercise of LLRP and followed the instructions of daily care (IDC) for a duration of 4-weeks. The participants in the CG were not involved in the rehabilitation protocols, but these participants only followed the IDC for a duration of 4-weeks.

### 2.6. Research Procedures

#### 2.6.1. Research Procedure of RPG

The researcher taught the strengthening exercises of LLRP and IDC to RPG for a duration of four weeks. Participants were advised to continue performing the strengthening exercises of LLRP three times a week for four weeks at home. These training sessions included strengthening exercises for the lower limbs in nonweight bearing, sitting, or lying positions. Each training session started with 10 minutes’ warm-up, 45–60 minutes of lower limb resistance training, and 10 minutes cool down at the end of the training protocol. A cool-down period is essential after a training session and should last approximately 5 – 10 minutes [23, 24]. When static stretching is used as part of a warm-up immediately prior to exercise, then it causes harm to muscle strength [25]. The participants in the RPG performed the strengthening exercises of LLRP and followed the IDC at home for four weeks. The feasibility, acceptability and contents of the IDC are explained in a recent randomized controlled trial [26].

The sequence of the training programme started with 10 minutes’ warm-up with whole body range of motion (ROM) and dynamic stretching exercises (Table 1). The participants performed 10 repetitions of ROM of each muscle group and five repetitions of dynamic stretching of each muscle group as a part of warm-up. After the warm-up, the participants performed the strengthening exercises of LLRP for the stipulated weeks as stated in Table 2. After completing the strengthening exercises, the participants performed the 10 minutes cool-down with whole body ROM and static stretching exercises (Table 1). The participants performed 10 repetitions of ROM of each muscle group and three repetitions of static stretching of each muscle group as a part of cool-down.

**Table 1:**
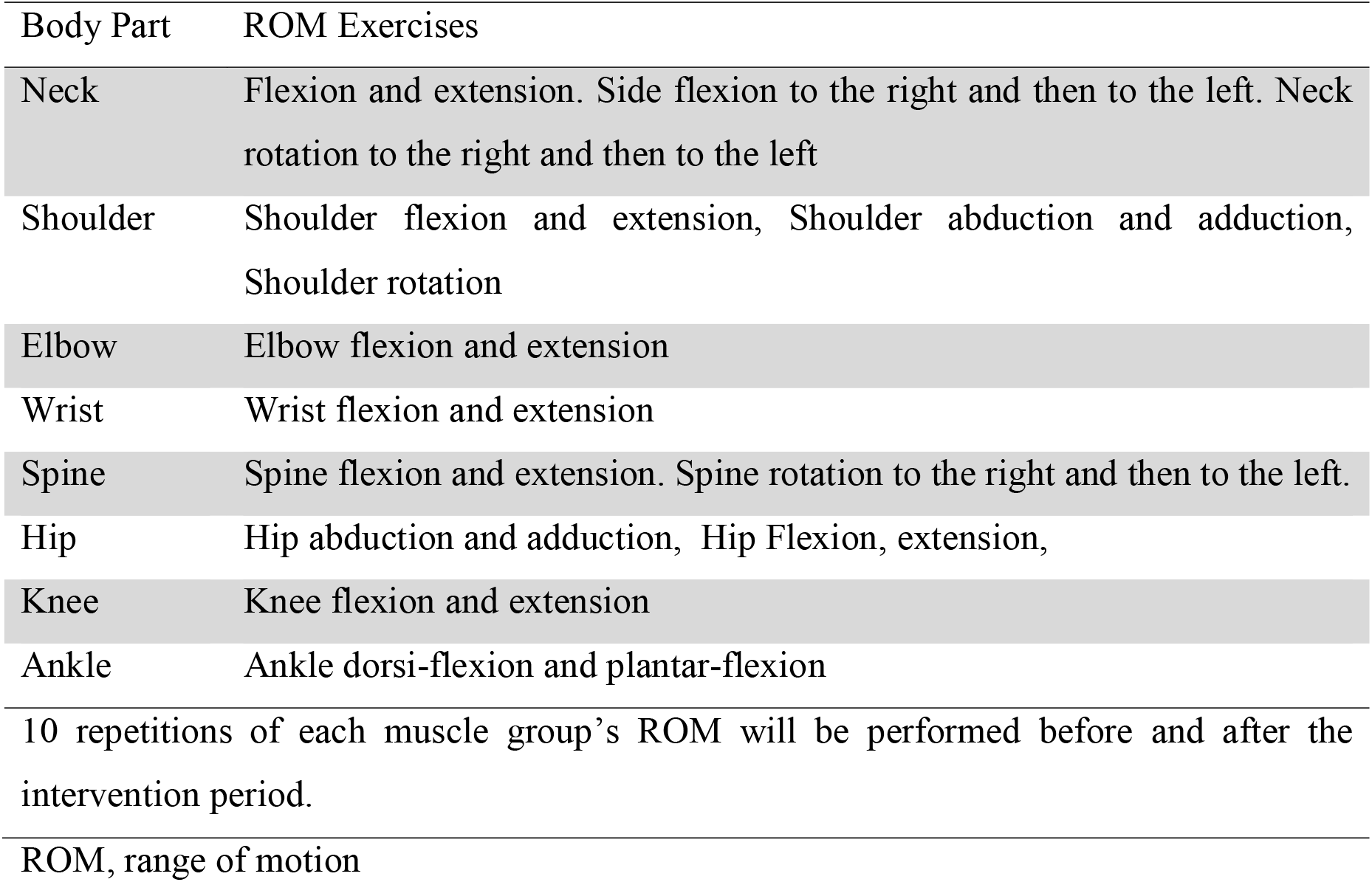
Whole body ROM exercises as part of warm-up or cool-down

**Table 2:** Strengthening exercises of LLRP in non-weight bearing sitting and lying positions

| Muscle group | Position | Resistance | For two weeks<br>(1<sup>st</sup> and 2<sup>nd</sup> week) | For two weeks<br>(3rd and 4th<br>week) |
| --- | --- | --- | --- | --- |
| Hip abductors | Supine<br>lying | Resistance<br>band | 2 sets of 7 reps | 2 sets of 10 reps |
| Hip adductors | Supine<br>lying | Resistance<br>band | 2 sets of 7 reps | 2 sets of 10 reps |
| Hip flexors | Side lying | Resistance<br>band | 2 sets of 7 reps | 2 sets of 10 reps |
| Hip extensors | Side lying | Resistance<br>band | 2 sets of 7 reps | 2 sets of 10 reps |
| Quadriceps<br>(Knee) | Side lying | Ankle weight | 2 sets of 7 reps | 2 sets of 10 reps |
| Hamstrings<br>(Knee) | Side lying | Ankle weight | 2 sets of 7 reps | 2 sets of 10 reps |
| Ankle dorsiflexors | Side lying | Foot weight | 2 sets of 7 reps | 2 sets of 10 reps |
| Ankle plantar<br>flexors | Side lying | Foot weight | 2 sets of 7 reps | 2 sets of 10 reps |

LLRP, lower limb rehabilitation protocol; reps, repetitions

#### 2.6.2. Research Procedure of CG

The participants in the CG were asked to follow the IDC only for a duration of four weeks. The IDC were also translated into Urdu language by two language experts as the participants’ the preference of the Urdu translation for better understanding based on a recent pilot study [27].

### 2.7. Outcomes Measures

The outcome measures were collected at baseline and after 4-weeks of intervention. Outcome measures were categorized into primary and secondary outcome measures.

#### 2.7.1. Primary Outcome Measures

These were knee pain, stiffness, and physical function assessed using the Western Ontario and McMaster Universities Osteoarthritis Index (WOMAC) score. The WOMAC score is widely accepted and validated [28]. The WOMAC score ranges from 0 to 4 on a Likert-type scale, the higher the score, the worse the pain, stiffness, and physical function.

#### 2.7.2. Secondary outcome measures

These were BMI, participant’s satisfaction, and exercise adherence. The BMI was calculated using the formula (weight (kilogram) / height (meter squared). Both the participant’s satisfaction and participant’s adherence to the interventions were assessed using a numeric rating scale ranging from 0 to 10.

Participants’ satisfaction with the RPG or CG intervention was determined by asking questions to know “How satisfied have you been with their leaflet provided intervention at home exercise program over the past 4-weeksã on a scale from zero = ‘not at all satisfied’ to 10 = ‘extremely satisfied. The numeric rating scale of a published study in which the participants were instructed to rate their satisfaction ranging from 0= ‘not at all satisfied’ to 10= ‘extremely satisfied was used [29]. Responses to the two interventions were analyzed separately.

Self-reported exercise adherence was measured by a numerical scale ranging from zero = never performed intervention of RPG or CG to 10 = always performed intervention of RPG or CG. Numerical rating scales from zero to 10 have good validity and reliability and have also been widely used in other trials [30, 31].

A study coordinator, who was blinded to the participant’s intervention, contacted all participants through a phone call upon study completion. The participants were asked about an opinion regarding the intervention of RPG or CG adherence. The blinded coordinator provided a score in response to the question ‘what is the score of 4-weeks RPG or CG intervention on a scale from zero = ‘never performed his intervention’ to 10 = ‘always performed the intervention according to their provided leaflets.

### 2.8. Statistical Procedures

Statistical Package for Social Sciences, version 22, Chicago, IL, was used to manage and analyze the data. Descriptive statistics was used for the demographic questionnaire and for the mean and standard deviation of all variables. Inferential statistics was used for all quantitative measures. Prior to data analysis, Shapiro-Wilk Test was used for all variables to check the normality of data. The scores were normally distributed; therefore, the paired samples t-test was used to analyze the differences of outcome measures within groups from baseline to post-test measurements. Analysis of variance 2 × 2 factor was used to compare the differences of clinical outcome measures between the groups. The independent sample t-test was used to evaluate the mean (95% confidence interval (CI) difference between groups for exercise adherence and participants’ satisfaction measured after 4-weeks of intervention.

## 3. Results

A total of seventy participants were screened and assessed for eligibility for inclusion in this study. Twenty participants were excluded for reasons as shown in Figure 1; the remaining fifty participants were randomized into RPG or CG. Of the twenty-five participants allocated to the RPG, 4 participants did not continue with their intervention because they were outstation due to occupation and sick. Likewise, 4 of the twenty-five participants allocated to the CG did not continue the intervention because they were travelling and unwilling. We could not obtain the postintervention outcomes for these 8 withdrawn participants. A final total of forty-two participants (twenty-one in the RPG and twenty-one in the CG completed the study and the data of which were analyzed (Figure 1).

**Figure 1.**
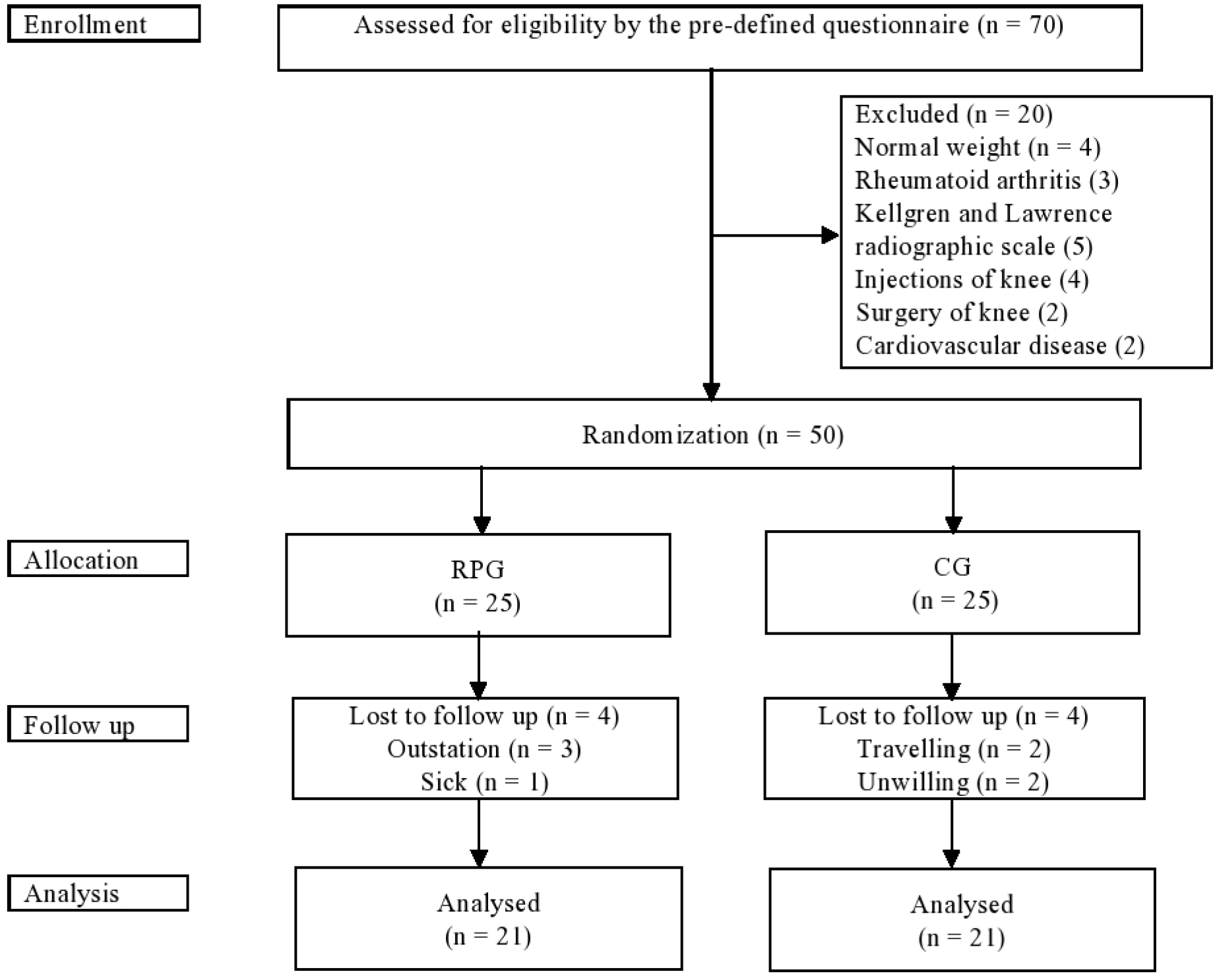
Flow chart of participants’ participation; RPG, Rehabilitation protocol group; CG, Control group; n, Number

Demographic characteristics and clinical outcome measures of the study participants at baseline are shown in Table 3. No significant differences were observed in baseline demographic characteristics and clinical outcome measures of pain, stiffness, physical function, and BMI between the 2 groups (p > 0.05).

**Table 3:** Baseline demographic characteristics and clinical outcome measures of participants at the baseline: mean (SD) or n (%)

| Baseline demographic characteristics and clinical outcome measures | Overall<br>(N = 50) | RPG<br>(n = 25) | CG<br>(n = 25) | p-value |
| --- | --- | --- | --- | --- |
| Age, mean (SD), y | 53.12 (5.41) | 53.40 (5.18) | 52.84 (5.74) | 0.719 |
| Gender (Male/Female), n | 23/27 | 11/14 | 12/13 | 0.782 |
| Educational Status, N (%) |  |  |  |  |
| • Matriculation | 8 (16) | 6 (24.0) | 2 (8.0) | 0.473 |
| • Intermediate | 20 (40) | 8 (32.0) | 12 (48.0) |  |
| • Bachelor | 13 (26) | 7 (28.0) | 6 (24.0) |  |
| • Master | 9 (18) | 4 (16.0) | 5 (20.0) |  |
| Employment, No. (%) |  |  |  |  |
| • Yes | 33 (66) | 19 (76.0) | 14 (56.0) | 0.141 |
| • No | 17 (34) | 6 (24.0) | 11 (44.0) |  |
| Weight (Kg), mean (SD) | 84.06 (11.09) | 86.68 (8.15) | 81.44 (13.05) | 0.095 |
| Height (m), mean (SD) | 2.63 (0.31) | 2.72 (0.34) | 2.54 (0.25) | 0.036 |
| BMI (kg/m<sup>2</sup>), mean (SD) | 32.09 (4.16) | 32.18 (4.49) | 32.01 (3.89) | 0.885 |
| WOMAC Pain (0-20), mean (SD) | 9.94 (1.73) | 9.84 (1.51) | 10.04 (1.94) | 0.687 |
| WOMAC Stiffness (0-8), mean (SD) | 3.70 (1.70) | 3.84 (1.90) | 3.56 (1.50) | 0.567 |
| WOMAC Physical function (0-68), mean (SD) | 23.60 (12.14) | 24.48 (12.77) | 22.72 (11.68) | 0.614 |

RPG, rehabilitation protocol group; CG, control group; SD, standard deviation; no, number; Kg, kilogram; m, meter

Table 4 shows that after 4-weeks of intervention, the participants in the RPG reported a significant reduction in pain (p ≤ 0.001) and stiffness (p ≤ 0.001), but no improvement in physical function (p = 0.104) and BMI (p = 0.364) within group. The participants in the CG also reported a reduction in pain, knee stiffness, physical function, and BMI scores in week 4 compared to baseline, the differences, however, were not statistically significant (p > 0.05) within group. Mean and 95% CI of outcome measures within group are shown in Figure 2.

**Table 4:**
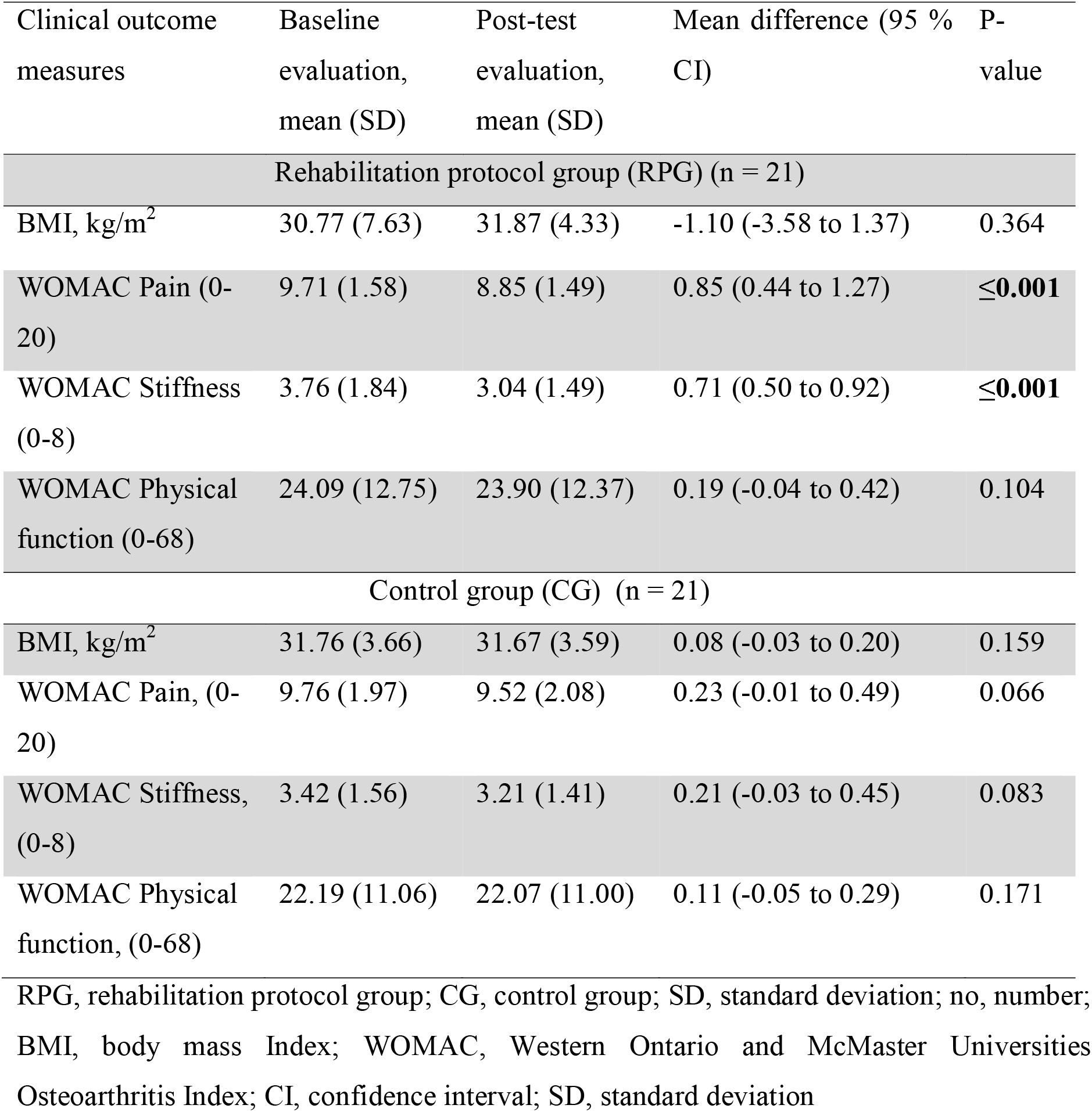
Clinical outcome measures of study participants within group

**Figure 2.**
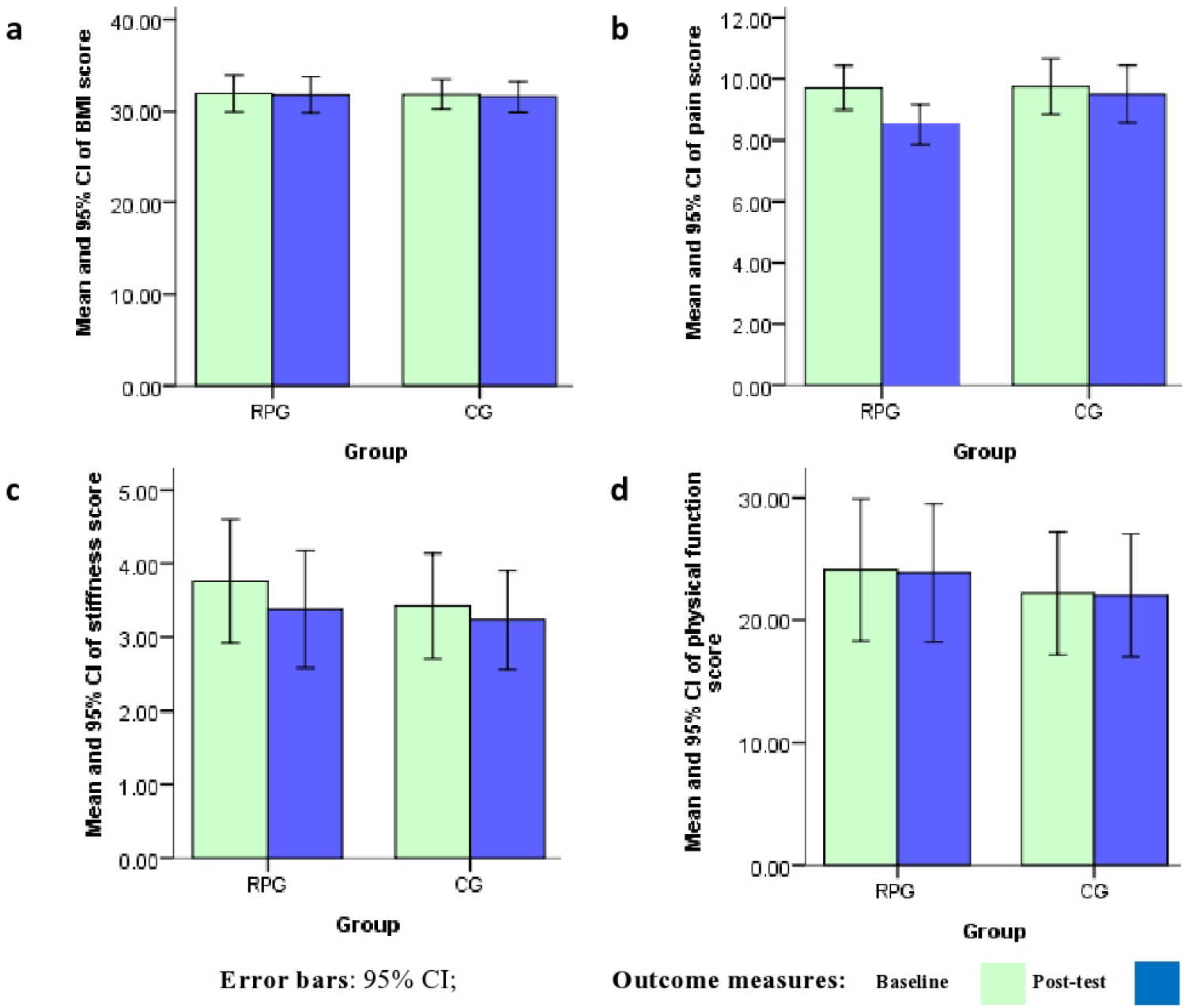
Mean and 95% CI of the outcomes measures within groups. **a** mean and 95% CI of BMI score at baseline and 3-month follow-up, **b** mean and 95% CI of pain score at baseline and 3-month follow-up, **c** mean and 95% CI of stiffness score at baseline and 3-month follow-up, **d** mean and 95% CI of physical function score at baseline and 3-month follow-up; RPG, Rehabilitation protocol group; CG, Control group; BMI, Body mass index; CI, Confidence interval

When the effectiveness of outcome measures was compared between the 2 groups, the participants in the RPG reported a statistically more significant improvement in the WOMAC pain (p = 0.016) and stiffness score (p = 0.002) than the CG. However, the participants in the RPG reported no statistically significant improvement in the scores of BMI and WOMAC physical function (p > 0.05) than the CG (Table 5).

**Table 5:** Comparison of clinical outcome measures between the groups (RPG and CG)

| Clinical outcome measures | RPG | CG | P-value |
| --- | --- | --- | --- |
|  | Mean difference (95<br>% CI) | Mean difference (95<br>% CI) |  |
| BMI, kg/m<sup>2</sup> | -1.10 (-3.58 to 1.37) | 0.08 (-0.03 to 0.20) | 0.868 |
| WOMAC Pain, (0-20) | 0.85 (0.44 to 1.27) | 0.23 (-0.01 to 0.49) | **0.016** |
| WOMAC stiffness, (0-8) | 0.71 (0.50 to 0.92) | 0.21 (-0.03 to 0.45) | **0.002** |
| WOMAC physical function, (0-68) | 0.11 (-0.05 to 0.29) | 0.11 (-0.05 to 0.29) | 0.576 |

RPG, rehabilitation protocol group; CG, control group; BMI, Body mass index; WOMAC, Western Ontario and McMaster Universities Osteoarthritis Index; CI, confidence interval

The mean between-group difference for participants’ satisfaction was 1.95 (95% CI 0.91 to 2.98) with a p-value of 0.001 in favour of the RPG. There was a statistically significant between-group difference for self-reported exercise adherence, with a mean between-group difference of 1.33 (95% CI 0.34 to 2.32) with a p-value of 0.010 in favour of the RPG. Similarly, there was a statistically significant between-group difference for coordinator-reported exercise adherence, with a mean between-group difference of 0.88 (95% CI 0.01 to 1.75) with a p-value of 0.046 in favour of the RPG (Table 6).

**Table 6:** Mean (SD) of groups and mean (95% CI) difference between groups for exercise adherence and participants’ satisfaction measured after 4 Weeks of interventions as post-test evaluation

| Outcomes | RPG | CG | RPG minus CG | P-value |
| --- | --- | --- | --- | --- |
| Self-reported exercise adherence (0 to 10) | 7.22 (1.47) | 5.88 (1.45) | 1.33 (0.34 to 2.32) | **0.010** |
| Coordinator-reported exercise adherence (0 to 10) | 7.02 (1.31) | 6.13 (1.25) | 0.88 (0.01 to 1.75) | **0.046** |
| Participants' satisfaction with the groups (0 to 10) | 8.92 (1.39) | 6.97 (1.66) | 1.95 (0.91 to 2.98) | **≤0.001** |

RPG, rehabilitation protocol group; CG, control group

There were no adverse as well as suspected unexpected serious adverse reactions reported in the current study.

## 4. Discussion

To the best of our knowledge, this is the first randomized controlled trial to address the short-term effects of strengthening exercise of the LLRP in improving pain, stiffness, physical function, BMI, patient, s satisfaction and exercise adherence among knee OA participants who are overweight or obese. Our results indicate that including the short-term effects of strengthening exercises in the LLRP could improve pain and stiffness more efficiently than could usual care. Similarly, the results of this study indicate that participants in the RPG reported greater satisfaction and adherence to their intervention compared to the participants in the CG.

The results for reducing pain of the current study are consistent with an overview of nine systematic reviews [32] and a current randomized controlled trial [33] that reported that exercise interventions for knee OA reduce pain but that effect sizes are considered small. In the current study, the strengthening exercises of LLRP were performed in nonweight bearing positions without putting mechanical pressure on the knee. Therefore, instead of a small duration, it reported a significant reduction in pain and stiffness. An overview of nine systematic reviews [32] and a randomized controlled trial [33] also reported significant improvement in physical function, but the current study’s results reported no improvement in physical function. It may be due to the small duration of the current study. The results of weight loss of the current study are inconsistent with a pragmatic randomized controlled trial that reported that telephone-based weight loss support did not affect weight [34]. A current study concluded that the progressive resistance strength training of LLRP in nonweight bearing positions in patients with knee OA is effective in reducing BMI [35].

A published study of short duration of 24 hours contradicts the current study and reported that a moderate strengthening exercise did not have an effect on knee pain, but it only induces a mild inflammatory response [36]. A study reported that an 18-month combined exercise and dietary weight-loss intervention of 316 overweight or obese individuals with radiographic evidence of knee OA was effective in improving knee pain as well as physical function [37]. It supports the current study’s results of knee pain. The results of a randomized controlled trial in which 90% participants were satisfied with their intervention of home exercise programs provided with an app with remote support [38], are the same as that of the current study. However, participants who received their intervention from home exercise programs with an app with remote support reported greater adherence [36] than that of the current study’s adherence to intervention. This current randomized controlled trial provides further evidence that the LLRP that include training sessions of strengthening exercises in nonweight bearing positions are more effective for overweight or obese knee OA participants than typical rehabilitation.

This study has several limitations. First, the results may not be generalized to all overweight or obese knee OA participants because we enrolled OA grading scale of 2-mild or 3-moderate according to Kellgren and Lawrence radiographic scale. No long-term follow-up records were taken. Finally, the comparisons were performed on a relatively smaller number of participants in this study. Thus, further research with a larger sample size and long-term follow-up are required to confirm the results of strengthening exercises of LLRP.

## 5. Conclusion

The current study showed the advantage of strengthening exercises of the LLRP in nonweight bearing positions on knee pain and stiffness reduction in overweight or obese participants with knee OA compared with IDC without strengthening exercises. Therefore, strengthening exercises of the LLRP in nonweight bearing positions may be an effective intervention to reduce knee pain and stiffness. The current study also showed that there was no improvement in physical function and BMI due to strengthening exercises of the LLRP that were performed for a duration of 4-weeks. In the management of overweight or obese participants with knee OA, strengthening exercises of major muscle groups of lower limbs in nonweight bearing positions may reduce knee pain and stiffness, and be an effective additional treatment option in the rehabilitation programme.

## Data Availability

All data produced in the present study are available upon reasonable request to the corresponding author

## Data Availability

The data used to support the findings of the study are available from the corresponding author upon request.

## Acknowledgments

The authors are so grateful to all participants in this study for their kindness and courage and for their valuable contribution to achieve the rehabilitation protocol.

## Supplementary Material

Additional File 1 - Trial Protocol

Additional File 2 - CONSORT 2010 Checklist

Additional File 3 - CONSORT-2010-Flow-Diagram

