## Supplementary Files for "Short-Term Effects of Strengthening Exercises of Lower Limb Rehabilitation Protocol on Pain, Stiffness, Physical Function and Body Mass Index among Knee Osteoarthritis Participants Who Were Overweight or Obese: A Clinical Trial"

### Clinical Trial Protocol
### Iranian Registry of Clinical Trials

18 Apr 2021

#### Short term effects of strengthening exercises of lower limb rehabilitation protocol (LLRP) on pain, stiffness, physical function and body mass index (BMI) among knee osteoarthritis (OA) participants who are overweight or obese.

##### Protocol summary

**Study aim**

To determine the short term effects of strengthening exercises of lower limb rehabilitation protocol on pain, stiffness, physical function and body mass index among knee osteoarthritis participants who are overweight or obese

**Design**

The study will be single blinded randomized controlled design.Three research coordinators will be involved in data collection. The one coordinator will be involved in baseline data collection and two coordinators will be involved in the assessment of outcomes. Participants will be blinded of other groups intervention.

**Settings and conduct**

The study will be conducted at the urban area of Lahore Pakistan and measurements will be assessed at the Teaching Bay of Rehmatul-Lil-Alameen Postgraduate Institute

**Participants/Inclusion and exclusion criteria**

Inclusion Criteria: Overweight or obese knee osteoarthritis participants, age between 45 and 60 years, residing in the urban community of Lahore, second and third degree of knee changes according to Kellgren and Lawrence radio-graphic scale Exclusion Criteria: System lupus erythematosus, Flat feet, Spinal deformities, Participants having history of metabolic, hormonal, orthopaedic and cardiovascular diseases, Previous surgery of knee/s of any cause The patients who had any injections of knee/s in the last six months or Unable to walk independently

**Intervention groups**

Intervention 1: Intervention group 1 the strengthening exercises of the lower limbs and instructions of daily care. The training session will be of 30-40 minutes. It will consist of three times a week for 4 weeks at their houses. Intervention 2: Intervention group 2 the participants in this group will follow the instructions of daily care only as a usual care for duration of 4 weeks at their houses.

**Main outcome variables**

Primary: Knee pain, stiffness and physical function
Secondary: Patients, s satisfaction and exercise adherence

##### General information

**Reason for update**

**Acronym**

**IRCT registration information**

IRCT registration number: **IRCT20191221045846N2**
Registration date: **2020-06-28, 1399/04/08**
Registration timing: **prospective**

Last update: **2020-06-28, 1399/04/08**
Update count: **0**

**Registration date**

2020-06-28, 1399/04/08

**Registrant information**


**Name**

Muhammad Tariq Rafiq

**Name of organization / entity**

Punjab Employees Social Security Institution

**Country**

Pakistan

**Phone**

+92 42 99330101

**Email address**


**Recruitment status**

**Recruitment complete**

**Funding source**

**Expected recruitment start date**
2020-07-10, 1399/04/20
**Expected recruitment end date**
2020-08-09, 1399/05/19
**Actual recruitment start date**
empty
**Actual recruitment end date**
empty
**Trial completion date**
empty

**Scientific title**
Short term effects of strengthening exercises of lower limb rehabilitation protocol (LLRP) on pain, stiffness, physical function and body mass index (BMI) among knee osteoarthritis (OA) participants who are overweight or obese.

**Public title**
Short term effects of exercises among knee osteoarthritis (OA) participants who are overweight or obese.
**Purpose**
Treatment
**Inclusion/Exclusion criteria**
**Inclusion criteria:**
Overweight or obese knee OA participants aged between 45 to 60 years. Familiar with WhatsApp application Participants will be chosen from the urban area community of Punjab Lahore, Pakistan. The sample included males and females having OA of one or both knees confirmed by medical specialist according to the Kellgren and Lawrence radiographic scale for the assessment of OA
**Exclusion criteria:**
Rheumatoid arthritis System lupus erythematosus Flat feet Spinal deformities Unable to walk independently History of metabolic, hormonal, orthopaedic or cardiovascular disease Previous surgery of knee/s Injections of knee/s for the last six months.
**Age**
From **45 years** old to **60 years** old
**Gender**
Both

**Phase**
2
**Groups that have been masked**

- Participant
- Outcome assessor
- Data analyser
- Data and Safety Monitoring Board

**Sample size**
Target sample size: **50**
**Randomization (investigator's opinion)**
Randomized
**Randomization description**
The researcher will allocate the selected knee osteoarthritis participants into two groups, namely, Rehabilitation Protocol Group (RPG) and Control Group (CG) by simple random technique using a computer-generated random number. The participants in the RPG will follow the strengthening exercise of LLRP and IDC for duration of 4-weeks. The participants in the CG will not involved in the rehabilitation protocols, but these participants will follow only the IDC for the duration of 4-weeks at their houses.
**Blinding (investigator's opinion)**
Single blinded
**Blinding description**
The principle investigator will not be blinded. The participants receiving the intervention will be blinded by simply not informing them of their treatment allocation. The coordinators collecting data will be independent individuals from trials and will be unaware of the group allocation. There will be different coordinators at the initial and final evaluation. Individuals performing the statistical analysis will be blinded by labelling the groups with non-identifying terms (such as X and Y).
**Placebo**
Not used
**Assignment**
Parallel
**Other design features**

#### Secondary Ids

empty

#### Ethics committees

**1**

**Ethics committee**
**Name of ethics committee**
Ethical Committee RAIC PESSI (Rehmatul-Lil-Alameen Postgraduate Institute of Cardiology, Punjab Empl
**Street address**
Multan Chongi Multan Road Lahore,
**City**
Lahore
**Postal code**
54000
**Approval date**
2020-05-20, 1399/02/31
**Ethics committee reference number**
RAIC PESSI/Estt/2020/36

#### Health conditions studied

**1**

**Description of health condition studied**
Osteoarthritis of knee
**ICD-10 code**
M17
**ICD-10 code description**
Osteoarthritis of knee

#### Primary outcomes

**1**

**Description**
knee pain

**Timepoint**
at baseline and after 4 weeks of intervention
**Method of measurement**
Western Ontario and McMaster Universities Osteoarthritis Index (WOMAC) score

**2**

**Description**
Stiffness
**Timepoint**
At baseline and after 4 weeks of intervention
**Method of measurement**
WOMAC score

**3**

**Description**
Physical function
**Timepoint**
At baseline and after 4 weeks of intervention
**Method of measurement**
WOMAC score

#### Secondary outcomes

**1**

**Description**
Body Mass Index
**Timepoint**
At baseline and after 4 weeks of intervention
**Method of measurement**
Calculated by the formula Weight (kg)/Height(meter square)

**2**

**Description**
Patients satisfaction
**Timepoint**
After 4 weeks of intervention
**Method of measurement**
Numeric rating scale ranging from 0 to 10

**3**

**Description**
Exercise adherence
**Timepoint**
After 4 weeks of intervention
**Method of measurement**
Numeric rating scale ranging from 0 to 10

#### Intervention groups

**1**

**Description**
The participants in this group (Rehabilitation Protocol Group) will perform the strengthening exercises of lower limb rehabilitation protocol (LLRP) and follow the instructions of daily care (IDC) for duration of four weeks at their houses according to the provided leaflet. Each training session of strengthening exercises will be performed three times a week for four weeks at their houses. These training sessions were the strengthening exercises of lower limbs in non-weight bearing sitting and lying positions. Each training session will consist of 45–60 minutes of training followed by 10 minutes warm up at the start and 10 minutes cool down at the end of training protocol.
**Category**
Treatment - Other

**2**

**Description**
The participants in this group (Control Group) will follow only the instructions of daily care (IDC) for duration of four weeks at their houses according to the provided leaflet.
**Category**
Treatment - Other

#### Recruitment centers

**1**

**Recruitment center**
**Name of recruitment center**
Teaching Bay of Rehmatul-Lil-Alameene Post Graduate Institute of Cardiology
**Full name of responsible person**
Dr Farid Ahmad Chaudhary
**Street address**
Teaching Bay of Rehmatul-Lil-Alameene Post Graduate Institute of Cardiology, Punjab Employees Social Security Institution, Multan Chongi, Multan Road Lahore
**City**
Lahore
**Postal code**
54000
**Phone**
+92 42 99330101
**Fax**
+92 42 99330098
**Email**


#### Sponsors / Funding sources

**1**

**Sponsor**
**Name of organization / entity**
Rehmatul-Lil-Alameene Post Graduate Institute of Cardiology
**Full name of responsible person**
Dr Farid Ahmad Chaudhary
**Street address**
Rehmatul-Lil-Alameene Post Graduate Institute of Cardiology, Punjab Employees Social Security

Institution, Multan Chongi, Multan Road Lahore
**City**
Lahore
**Postal code**
54000
**Phone**
+92 42 99330101
**Fax**
**Email**

**Grant name**
**Grant code / Reference number**
**Is the source of funding the same sponsor organization/entity?**
Yes
**Title of funding source**
Rehmatul-Lil-Alameene Post Graduate Institute of Cardiology
**Proportion provided by this source**
50
**Public or private sector**
Public
**Domestic or foreign origin**
Domestic
**Category of foreign source of funding**
*empty*
**Country of origin**
**Type of organization providing the funding**
Other

**2**

**Sponsor**
**Name of organization / entity**
University of Malaya
**Full name of responsible person**
Dr Eliza Hafiz
**Street address**
Centre for Sport and Exercise Sciences, University of Malaya, Kuala Lumpur, Malaysia
**City**
Kuala Lumpur
**Postal code**
42000
**Phone**
+60 3-7967 3327
**Email**

**Grant name**
**Grant code / Reference number**
**Is the source of funding the same sponsor organization/entity?**
No
**Title of funding source**
N/A
**Proportion provided by this source**
50
**Public or private sector**
Public
**Domestic or foreign origin**
Foreign
**Category of foreign source of funding**
Sponsor: country of origin
**Country of origin**
PK
**Type of organization providing the funding**
Other

#### Person responsible for general inquiries

**Contact**
**Name of organization / entity**
Punjab Employees Social Security Institution
**Full name of responsible person**
Muhammad Tariq Rafiq
**Position**
Consultant Senior Physiotherapist
**Latest degree**
Medical doctor
**Other areas of specialty/work**
Physiotherapy
**Street address**
54000 B. Rehmatul-Lil-Alamine Post Graduate Institute of Cardiology, Punjab Employees Social Security Institution, Multan Chongi, Multan Road Lahore
**City**
Lahore
**Province**
Punjab
**Postal code**
54000
**Phone**
+92 42 99330101
**Fax**
**Email**


#### Person responsible for scientific inquiries

**Contact**
**Name of organization / entity**
University of Malaya
**Full name of responsible person**
Mohamad Shariff A Hamid
**Position**
Associate Professor
**Latest degree**
Ph.D.
**Other areas of specialty/work**
Physical Medicine
**Street address**
Sports Medicine Department, University of Malaya Medical Centre, Kuala Lumpur, Malaysia
**City**
Kuala Lumpur
**Province**
Selangor
**Postal code**
59100
**Phone**
+60 3-7967 6669
**Email**


#### Person responsible for updating data

**Contact**

**Name of organization / entity**
Punjab Employees Social Security Institution

**Full name of responsible person**
Muhammad Tariq Rafiq

**Position**
Consultant Senior Physiotherapist

**Latest degree**
Medical doctor

**Other areas of specialty/work**
Physiotherapy

**Street address**
54000 B. Rehmatul-Lil-Alamine Post Graduate Institute of Cardiology, Punjab Employees Social Security Institution, Multan Chongi, Multan Road Lahore

**City**
Lahore

**Province**
Punjab

**Postal code**
54000

**Phone**
+92 42 99330101

**Fax**

**Email**


#### Sharing plan

**Deidentified Individual Participant Data Set (IPD)**
Undecided - It is not yet known if there will be a plan to make this available

**Study Protocol**
Undecided - It is not yet known if there will be a plan to make this available

**Statistical Analysis Plan**
Undecided - It is not yet known if there will be a plan to make this available

**Informed Consent Form**
Undecided - It is not yet known if there will be a plan to make this available

**Clinical Study Report**
Undecided - It is not yet known if there will be a plan to make this available

**Analytic Code**
Undecided - It is not yet known if there will be a plan to make this available

**Data Dictionary**
Undecided - It is not yet known if there will be a plan to make this available
