## Supplementary Files for "Short-Term Effects of Strengthening Exercises of Lower Limb Rehabilitation Protocol on Pain, Stiffness, Physical Function and Body Mass Index among Knee Osteoarthritis Participants Who Were Overweight or Obese: A Clinical Trial"

**CONSORT 2010 Flow Diagram**

**Allocation**

**Analysis**

**Follow-Up**

**Enrollment**

Assessed for eligibility (n= )

Excluded (n=	)

♦  Not meeting inclusion criteria (n= )

♦  Declined to participate (n= )

♦  Other reasons (n= )

Analysed (n= )
♦ Excluded from analysis (give reasons) (n= )

Lost to follow-up (give reasons) (n= )

Discontinued intervention (give reasons) (n= )

Allocated to intervention (n= )

♦ Received allocated intervention (n= )

♦ Did not receive allocated intervention (give reasons) (n= )

Lost to follow-up (give reasons) (n= )

Discontinued intervention (give reasons) (n= )

Allocated to intervention (n= )

♦ Received allocated intervention (n= )

♦ Did not receive allocated intervention (give reasons) (n= )

Analysed (n= )
♦ Excluded from analysis (give reasons) (n= )

Randomized (n= )
